# Digital exclusion and mental health in UK Armed Forces veterans: findings from the Veterans Digital Needs Study

**DOI:** 10.64898/2026.06.22.26356243

**Authors:** Daniel Leightley, Emily Gillings, Pippa Boering, Kathryn Dalrymple, Vasa Curcin, Neil Greenberg, Charlotte Williamson

## Abstract

**Background:** Public services are increasingly delivered through digital platforms. Although digital health may improve access and scalability, they may also widen inequalities for people who lack reliable access, confidence, skills, affordability or trust.

**Objective:** This study examined the prevalence of self-reported digital exclusion among UK veterans and assessed its association with depression, anxiety and loneliness.

**Methods:** A cross-sectional online survey was conducted between July 2025 and March 2026. Participants were UK Armed Forces veterans and resident in the UK. The survey collected sociodemographic, military service, digital access and health data. Self-reported digital exclusion was defined as reporting feeling excluded or disadvantaged due to lack of digital access or skills. Probable depression, anxiety and loneliness were assessed using the PHQ-2, GAD-2 and three-item UCLA Loneliness Scale, respectively. Associations between digital exclusion and each outcome were examined using adjusted multivariable logistic regression.

**Results:** Of 1,911 responses received, 1,607 were included after data quality exclusions. Among participants with valid responses to the primary digital exclusion item, 553 (41.7%) reported digital exclusion. Digital exclusion was more common among females, younger veterans and those with lower household income. Probable depression, anxiety and loneliness were more prevalent among digitally excluded participants than among non-excluded participants. In adjusted models, self-reported digital exclusion was associated with higher odds of probable depression (AOR 1.38; 95% CI 1.04 to 1.83; *p*=0.028), probable anxiety (AOR 1.63, 95% CI 1.23 to 2.16; *p*<0.001), and probable loneliness (AOR 1.85; 95% CI 1.43 to 2.40; *p*<0.001).

**Conclusion:** More than two-fifths of veterans with valid exposure data reported digital exclusion, despite high reported device access and confidence. Self-reported digital exclusion was associated with poorer mental health and loneliness, although causality cannot be inferred from these cross-sectional data. Digital-first services for veterans should include routine digital needs screening, targeted support and clear non-digital routes to care.

## Introduction

In the United Kingdom (UK), health and public services are being transformed by digital technologies and generative artificial intelligence (GenAI). In 2025, the UK government introduced a ten-year plan to make the NHS ‘digital first’, and the NHS App the ‘front door’ to services [1]. Digital therapeutics, such as smartphone-delivered apps for hazardous alcohol consumption or PTSD, have the potential for personalised, scalable interventions for veterans and other groups [2–4]. However, digital innovation also presents risks to certain populations; if patients lack reliable internet access, appropriate devices, or the skills and confidence to navigate digital systems, they may be unable to benefit [5]. Digital exclusion refers to deficiencies in access, affordability, skills or trust. It is not a binary state but a spectrum; while some individuals have no internet access, live in rural areas, others have devices but lack the ability or confidence to use them effectively. Even when access is available, low digital literacy, educational attainment, cognitive impairment or language barriers can prevent meaningful use, thereby widening inequalities [6].

The scale of digital exclusion in the UK is significant and widespread. Approximately 4.5 million people have never been online, of this group 94% are aged over 55, and an estimated 6% of households have no internet access with rural communities disproportionately affected [7]. Research has shown that digital skills deficits are more common among people with disabilities and those with low income, and roughly one-third of people over the age of 75 lack basic digital skills [7]. Research has shown that age, socioeconomic status, disability and geography intersect with digital exclusion, and that an individual’s position on the continuum can change over time [8]. As health services transition to the new ‘digital first’ approach, this spectrum of exclusion will intensify existing health inequalities.

Emerging evidence links digital exclusion to poorer health and social isolation. A longitudinal analysis of older adults across 24 countries found that digital exclusion was associated with a higher incidence of depressive symptoms even after adjustment for household wealth (adjusted incidence ranging from 1.30 to 1.62 across cohorts). The association was stronger among individuals in lower-wealth households and, in most cohorts, among those who did not regularly interact with their children (after adjusting for household wealth) [9]. Qualitative research with people living with severe mental illness has described how digital exclusion exacerbates loneliness, undermines self-efficacy and limits access to health and social services. Conversely, digital health interventions offer opportunities to reduce barriers and improve outcomes, but these benefits depend on inclusive design and targeted support [10].

Veterans constitute a sizeable and distinctive population in the UK. Approximately 150,000 personnel currently serve in the UK Armed Forces, while around 1.85 million people in England and Wales are veterans [11,12]. Service experiences, such as deployments, combat exposure and military culture, shape veterans’ health and wellbeing [13]. Some veterans face challenges during transition to civilian life, including physical and mental health conditions, social isolation and employment difficulties [14,15]. Digital innovations have a lot of opportunity for veterans, with tailored apps and telehealth services emerging to support mental health, substance use and chronic disease management [16–19]. Yet digital inclusion among veterans cannot be assumed.

A recent large-scale quality-improvement initiative across 14 Veterans Affairs (VA) medical centres in the United States (US) integrated digital needs screening and found that 42.7% of veterans reported at least one digital need [20]. Digital needs were more prevalent among older veterans, especially those aged 65 years and above; and, more common among Black or African American veterans and those with greater clinical complexity. Despite increasing recognition of digital inequalities, there is a dearth of research on how digital exclusion relates to mental health and loneliness in UK veterans. This study examines associations between self-reported digital exclusion and screening positive for probable depression, probable anxiety, and probable loneliness among UK Armed Forces veterans. This is the first study of its kind in the UK focusing on the UK Armed Forces. We hypothesised that veterans reporting digital exclusion would have higher odds of meeting screening thresholds for these outcomes. We also described patterns of digital access, cost and privacy barriers, and how digital exclusion varied across key sociodemographic and service-related sub-groups.

## Methods

Ethical approval for this study was obtained from King’s College London Research Ethics Committee (MRA-24/25-47812). All participants provided informed consent prior to providing study data.

### Study design and setting

This study was a cross-sectional online survey of digital needs among UK Armed Forces veterans. Data were collected between July 2025 and March 2026 using an anonymous Qualtrics questionnaire designed to take approximately 10 to 15 minutes to complete. Survey content covered sociodemographic characteristics, military service background, health status, digital access, digital barriers and digital service use. Items were adapted from a previously implemented United States veterans’ digital needs methodology to support comparability, with modifications to reflect UK language, culture and service context [20].

Unlike the US study, which gathered information via clinical screening within the VA health system, the present study relied on self-enrolment through an online survey link which was promoted through a range of communication channels including social media, print media, email lists and networks [21,22]. This approach was adopted because UK veterans are not uniformly registered within a single, dedicated veteran healthcare system; therefore, broad online recruitment routes were used. Further, veteran status is not routinely collected in the UK healthcare system.

### Participants

The target population comprised UK Armed Forces veterans, defined as individuals who had served at least one paid day in the UK Armed Forces, including former Regular and Reserve personnel. Inclusion criteria were: (1) self-reported veteran status; (2) aged 18 years or older; (3) resident in the UK; and (4) able to provide informed consent and complete the questionnaire in English. Veterans were not excluded based on discharge date, length of service beyond the minimum veteran definition, or reason for leaving service, to capture a broad range of experiences from recent service leavers to older veterans. Exclusion criteria were: (1) current serving members of the UK Armed Forces; (2) inability to complete the questionnaire in English; and (3) failure to provide informed consent.

### Measures

All items on the questionnaire were optional (Table 1). Sociodemographic and military service characteristics were captured to describe the sample and for use as covariates in analyses.

**Table 1:**
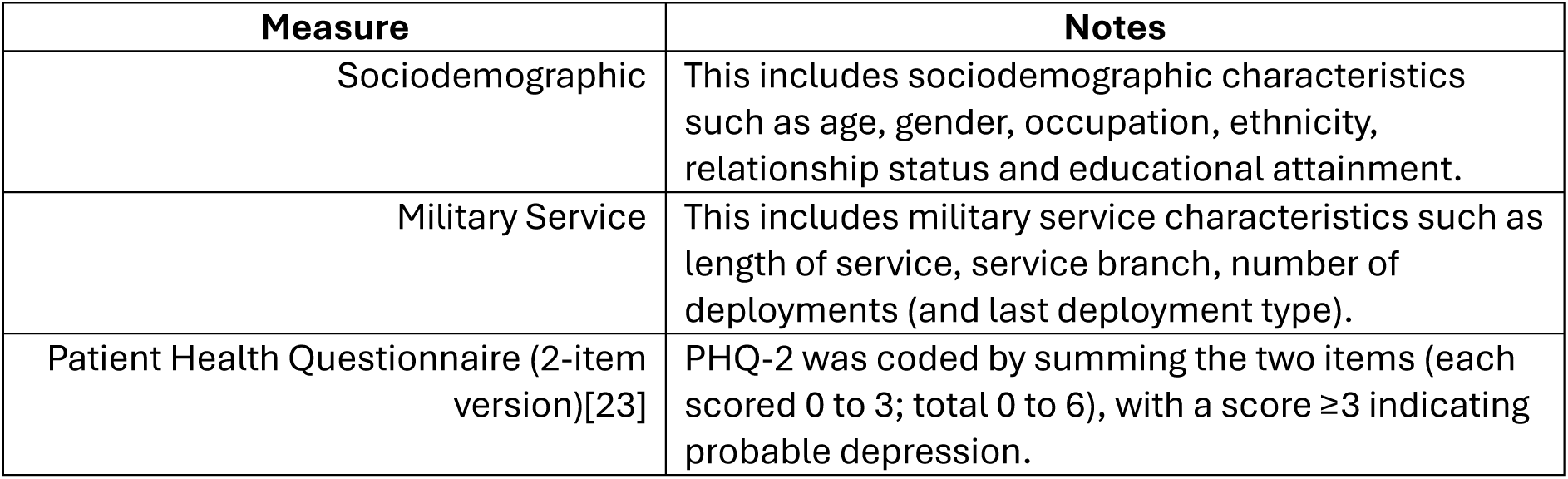
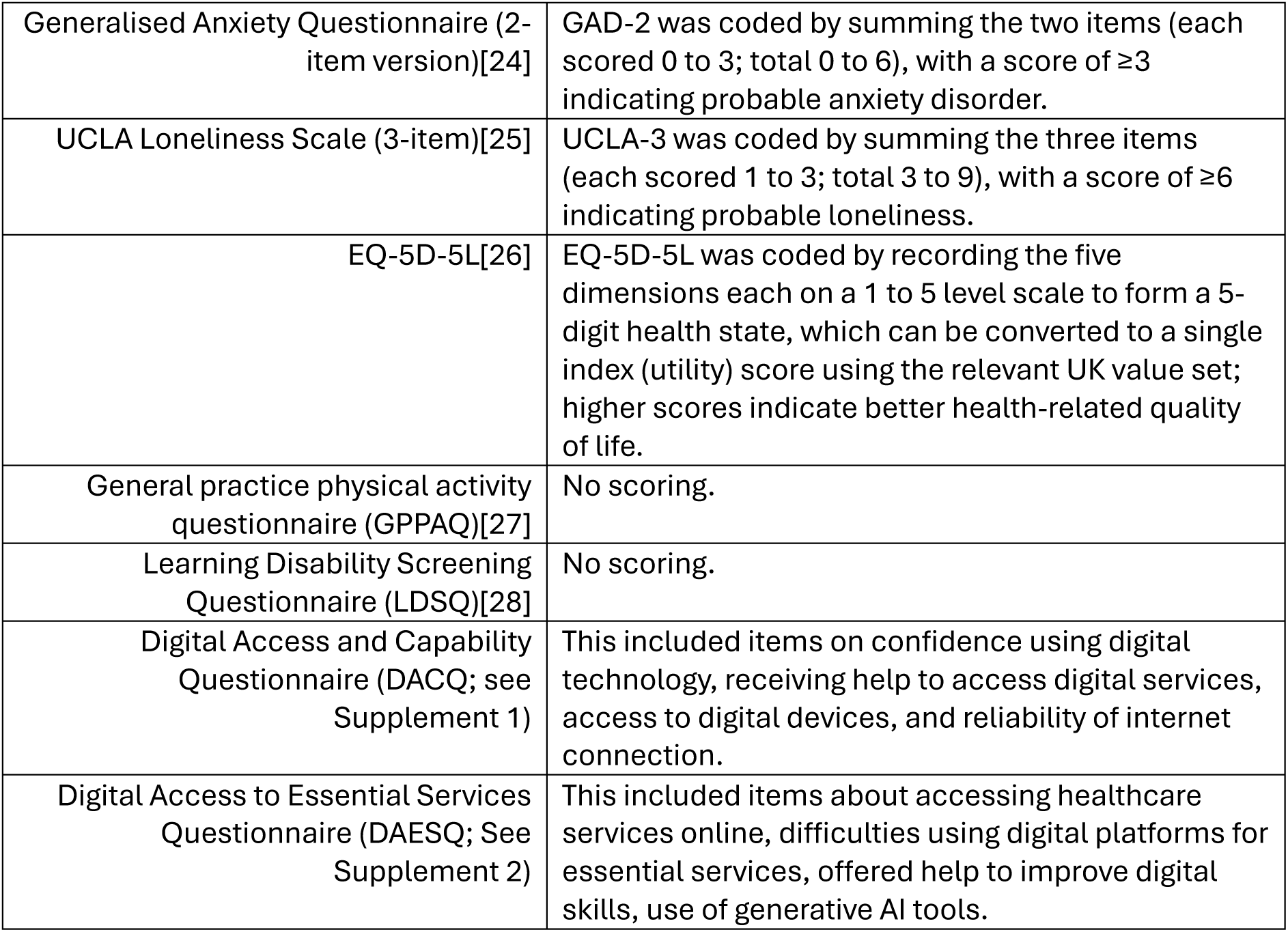
Questionnaires used in the Veterans’ Digital Needs survey.

#### Mental health outcomes

Depressive symptoms, anxiety symptoms, and loneliness were measured using the PHQ-2 [23], GAD-2 [24], and UCLA-3 [25], respectively.

#### Health and wellbeing

Health-related quality of life (EQ-5D-5L; [26]), physical activity (GPPAQ; [27]), and learning disability screening (LDSQ; [28]) were included to characterise the sample and to support interpretation of digital exclusion patterns across health and functioning.

#### Digital access, barriers, and service use

Digital access and capability were assessed using the DACQ, and use of online services was assessed using the Digital Access to Essential Services Questionnaire (DAESQ). These were developed for this study and are available for use.

#### Primary exposure

Digital exclusion was defined using the item “Have you ever felt excluded or disadvantaged due to lack of digital access or skills?” As part of the DAESQ. This measure should be interpreted as self-reported digital exclusion, capturing perceived disadvantage related to digital access, skills or participation rather than objective absence of internet access or digital devices. Participants responding “Yes, often” or “Yes, occasionally” were classified as digitally excluded; those responding “No” were classified as not digitally excluded. “Not sure” responses were treated as missing for this indicator.

### Statistical analysis

Analyses were conducted in Stata version 17.0. Prior to analysis, responses were screened for data quality and potential duplicate or low-quality entries. Responses were excluded if they were flagged as duplicate responses, had a Qualtrics Fraud Risk score greater than 0.5, or were completed in less than 5 minutes. The survey was designed to take approximately 10 to 15 minutes to complete; therefore, completion in less than 5 minutes was considered implausibly rapid for meaningful completion of the consent process and questionnaire items.

Descriptive statistics were used to summarise participant characteristics, digital access, digital barriers and outcome caseness. Continuous variables were summarised using means and standard deviations, and categorical variables were summarised using frequencies and percentages. Denominators varied across analyses because questionnaire items were optional and “not sure” responses were treated as missing where specified. For the primary exposure, participants responding “Yes, often” or “Yes, occasionally” to the digital exclusion item were classified as reporting self-reported digital exclusion, while those responding “No” were classified as not reporting digital exclusion. Participants responding “Not sure” were excluded from analyses involving the primary exposure.

The primary outcomes were binary caseness indicators for probable depression, probable anxiety and probable loneliness. Probable depression was defined as PHQ-2 score ≥3, probable anxiety as GAD-2 score ≥3, and probable loneliness as UCLA-3 score ≥6. Outcome prevalence was first summarised overall and stratified by self-reported digital exclusion.

Associations between self-reported digital exclusion and each outcome were examined using separate multivariable logistic regression models. Models adjusted for age, gender, educational attainment, household income, disability, UK region, military service branch, service type, rank group and number of deployments. Robust standard errors were used. Regression models were fitted using complete-case data for the outcome, exposure and covariates included in each model.

Results are presented as adjusted odds ratios (AOR) with 95% confidence intervals and *p*-values. To aid interpretation, adjusted predicted probabilities were estimated using post-estimation margins. Adjusted probability differences were calculated as the predicted probability among participants reporting digital exclusion minus the predicted probability among participants not reporting digital exclusion and are presented in percentage points. Study findings are reported following the STROBE Checklist for cross-sectional studies [29].

## Results

### Sample characteristics

In total 1,911 responses were received, with 198 excluded due to completion within 5 minutes, 84 responses excluded due to duplication and 22 responses excluded based on Qualtrics Fraud Risk score >0.5. A total of 1,607 UK Armed Forces veterans completed the survey and were included in the descriptive analyses (see Table 2). Mean age was 43.7 years (SD 15.7; *n*=1,597), with *n*=591 (36.8%) aged <35 years. Participants were predominantly male (*n*=1,046, 65.1%). Educational attainment was distributed across GCSE or below/other (*n*=512, 31.9%), A-level or equivalent (*n*=510, 31.7%), and degree or above (*n*=576, 35.8%).

**Table 2:** Sample characteristics (n=1C07).

| Characteristic | *n* | % |
| --- | --- | --- |
| Age (years) |  |  |
| <35 | 591 | 36.8 |
| 35-44 | 439 | 27.3 |
| 45-54 | 141 | 8.8 |
| 55-64 | 181 | 11.3 |
| 65+ | 245 | 15.3 |
| Gender |  |  |
| Male | 1,046 | 65.1 |
| Female | 552 | 34.4 |
| Educational attainment |  |  |
| GCSE or below/other | 512 | 31.9 |
| A-level or equivalent | 510 | 31.7 |
| Degree or above | 576 | 35.8 |
| Household income |  |  |
| <£15,000 | 55 | 3.4 |
| £15,000-£23,999 | 373 | 23.2 |
| £24,000-£39,999 | 460 | 28.6 |
| £40,000-£54,999 | 445 | 27.7 |
| >£55,000 | 265 | 16.5 |
| Any disability | 272 | 16.9 |
| Service branch |  |  |
| Army and Royal Marines | 1,033 | 64.3 |
| Royal Navy | 377 | 23.5 |
| RAF | 179 | 11.1 |
| Rank |  |  |
| Junior ranks | 947 | 58.9 |
| Senior ranks | 445 | 27.7 |
| Officer ranks | 156 | 9.7 |
| Other or unspecified | 41 | 2.6 |

Percentages may not sum to 100% due to missingness. Rank structures were combined due to the way the questions were presented.

Household income was most reported in the £24,000 to £39,999 (*n*=460, 28.6%) and £40,000 to £54,999 (*n*=445, 27.7%) ranges. Overall, *n*=272 (16.9%) reported a disability. Most participants had served in the Army and Royal Marines (*n*=1,033, 64.3%), with the majority being from the junior ranks (*n*=947, 58.9%), and *n*=445 (27.7%) from senior ranks (which includes Senior Non-Commissioned such as Warrant Officer).

### Digital access, barriers, and digital exclusion

Most participants reported access to multiple device types (see Table 3); *n*=999 (62.2%) reported access to three device types and *n*=472 (29.4%) reported access to four or more device types. Despite generally high reported confidence using digital technologies (*n*=1339, 83.3%, “very confident”), cost constraints were common (*n*=967, 60.2%). Privacy concerns were reported by *n*=417 (26.0%), with a “not sure” response for this item (*n*=391, 24.3%). Among participants with valid responses to the primary digital exclusion item, 553/1,327 (41.7%) met criteria for digital exclusion.

**Table 3:** Digital access and barriers (n=1C07).

| Measure | n | %* |
| --- | --- | --- |
| Device access count: |  |  |
| 0 devices | 29 | 1.8 |
| 1-2 device types | 107 | 6.6 |
| 3 device types | 999 | 62.2 |
| 4+ device types | 472 | 29.4 |
| Reported 1+ difficult digital tasks | 149 | 9.3 |
| Runs out of phone minutes/data sometimes or often | 928 | 57.7 |
| Very confident using digital technologies | 1,339 | 83.3 |
| Training/support in last 12 months | 903 | 56.2 |
| Cost barrier (yes) | 967 | 60.2 |
| Privacy barrier (yes) | 417 | 26.0 |
| Accessed healthcare services online in past 12 months (yes) | 839 | 52.2 |
| Accessed government/public services online in past 12 months (yes) | 783 | 48.7 |
| Ever experienced difficulties using digital platforms for essential services (yes) | 423 | 26.3 |

\*Percentages may not sum to 100% due to missingness.

Digital exclusion differed across participant subgroups (see Table 4). Digital exclusion was more common among females than males (53.5% compared to 35.9%) and was highest in younger age groups (<35: 60.3%; 35 to 44: 51.4%). Digital exclusion was lowest among those reporting household income >£55,000 (11.1%).

**Table 4:** Digital exclusion by key demographics.

| Subgroup | Digital exclusion yes (total)* | % |
| --- | --- | --- |
| Gender male | 321/894 | 35.9 |
| Gender female | 231/432 | 53.5 |
| Age <35 | 270/448 | 60.3 |
| Age 35-44 | 182/354 | 51.4 |
| Age 45-54 | 16/130 | 12.3 |
| Age 55-64 | 27/172 | 15.7 |
| Age 65+ | 58/223 | 26.0 |
| Income £15,000-£23,999 | 165/297 | 55.6 |
| Income £24,000-£39,999 | 168/367 | 45.8 |
| Income £40,000-£54,999 | 173/364 | 47.5 |
| Income >£55,000 | 28/252 | 11.1 |
| Disability yes | 68/250 | 27.2 |
| Service type regular/full-time reserve | 545/1,260 | 43.3 |
| Service type reserve/other | 8/66 | 12.1 |
| Rank junior ranks | 355/777 | 45.7 |
| Rank senior ranks (NCO/WO) | 156/363 | 43.0 |

\*Counts may not match due to missingness.

### Mental health and loneliness outcomes

Across all three outcomes, probable symptoms were more common among veterans reporting self-reported digital exclusion than among those not reporting digital exclusion (see **Error! Not a valid bookmark self-reference.**). In unadjusted comparisons, PHQ-2 caseness was 56.7% among digitally excluded participants compared with 29.0% among non-excluded participants. Similar differences were observed for GAD-2 caseness (57.9% compared with 29.1%) and UCLA-3 loneliness caseness (63.9% compared with 42.7%).

In adjusted logistic regression models, self-reported digital exclusion remained associated with higher odds of probable depression, probable anxiety and probable loneliness. The adjusted odds ratio was 1.38 for PHQ-2 caseness, 1.63 for GAD-2 caseness and 1.85 for UCLA-3 caseness. Adjusted predicted probabilities were also higher among digitally excluded veterans across all outcomes, with adjusted absolute differences of 5.8 percentage points for probable depression, 9.3 percentage points for probable anxiety and 13.8 percentage points for probable loneliness.

**Table 5:** Outcome caseness and adjusted associations between self-reported digital exclusion and mental health/loneliness outcomes.

| Outcome | Crude caseness, non-excluded vs excluded | AOR (95% CI; *p* value) | Adjusted probability difference, percentage points |
| --- | --- | --- | --- |
| PHQ-2 caseness (≥3) | 29.0% vs 56.7% | 1.38 (1.04 to 1.83; *p*=0.028) | +5.8 |
| GAD-2 caseness (≥3) | 29.1% vs 57.9% | 1.63 (1.23 to 2.16; *p*<.001) | +9.3 |
| UCLA-3 caseness (≥6) | 42.7% vs 63.9% | 1.85 (1.43 to 2.40; *p*<.001) | +13.8 |

## Discussion

This study provides the first quantitative picture of digital exclusion and its relationship with mental health in UK Armed Forces veterans. Despite high reported access to multiple devices and high self-rated confidence, more than two-fifths of participants reported feeling excluded or disadvantaged due to lack of digital access or skills. Digital exclusion was not merely an absence of connectivity; almost all participants had some form of internet and device access yet still felt excluded. This highlights a divide in the use and utility of technology rather than purely in access. Veterans who were digitally excluded were much more likely to meet screening thresholds for depression, anxiety and loneliness. Digital exclusion was associated with higher odds of all probable adverse mental health outcomes, namely depression, anxiety disorder, and loneliness. These associations persisted after adjusting for known confounders, suggesting that digital exclusion may independently contribute to poorer mental health and social isolation among UK veterans.

The association between digital exclusion and poorer mental health aligns with a growing literature linking digital access, skills and confidence with psychological outcomes. However, the direction of this association cannot be established from our cross-sectional data. Digital exclusion may limit access to services, support and social connection, but poorer mental health may also make it harder for individuals to engage confidently with digital platforms. Symptoms of depression or anxiety may reduce motivation, concentration and perceived self-efficacy, while loneliness may reduce opportunities to receive informal digital support. These findings should therefore be interpreted as evidence of an association, not causation. For example, a longitudinal study across 24 countries found that older adults who were digitally excluded had a higher incidence of depressive symptoms; this relationship was strongest among individuals with low wealth or infrequent contact with their children [9].

Digital exclusion has been conceptualised as a three-level digital divide [30]: 1) the access divide (lack of connectivity or devices), 2) the use divide (differences in ability to use technology effectively), and 3) the utility divide (differences in deriving social or economic benefits from technology). Early research on the digital divide has shown that disparities exist both within countries, shaped by socioeconomic status, geography, education, and disability, and between higher- and lower-income settings. It has also emphasised the need for coordinated responses that address both infrastructure and digital skills, including the potential role of trusted community institutions [31]. Further, a consideration around adapting digital tools to make them more inclusive and easier should be considered. Recent evidence reinforces this multidimensional view by showing an inequality loop, where socially and economically vulnerable groups may be online yet still lack the experience needed to gain benefits from digital participation, thereby reinforcing their existing disadvantage [32]. Our study population reported high levels of access, but persistent use and utility divides may explain why a substantial proportion reported digital exclusion and poorer mental health.

Several interrelated mechanisms may link digital exclusion to depressive symptoms, anxiety and loneliness. First, cost barriers and limited data or device credit was common in our sample. Financial stress related to paying for internet and data is a recognised driver of digital exclusion and can independently contribute to mental distress [33]. Second, many participants reported difficulty in completing online tasks or a lack of confidence using digital technology. Low digital literacy and computer-related anxiety are widely reported barriers to engaging with digital health and social interventions [34]. Research in people living with severe mental illness shows that digital exclusion is often due to a lack of knowledge or skills, inability to access technology because of personal circumstances, and barriers related to mental health symptoms [35]. Such deficits may foster frustration, reduce self-efficacy/agency and deter individuals from seeking help or connecting with others online. Third, digital exclusion may exacerbate social isolation.

People with severe mental illness who lack internet access or skills report higher levels of loneliness and less access to benefits, employment and support services [35]. Even among those with access, limited- or non-use is common; a study during the Covid-19 pandemic found people with severe mental ill health were limited- or non-users despite having internet access, often due to low motivation, difficulty using technology and security concerns [36]. Similar patterns may apply to veterans, particularly those with long-term physical and mental health conditions.

The fact that younger veterans (<35 years) and females in our sample were more likely to report digital exclusion suggests that digital inequality is not confined to older adults. Evidence suggests that men, older individuals and those with psychosis are most at risk of digital disengagement [37]. However, digital skills gaps may intersect with other forms of marginalisation. For example, younger veterans may lack digital financial resources or live in precarious housing where connectivity is unreliable. Women in the general population are less likely to engage with certain forms of technology and may have different privacy concerns [38]. The higher digital exclusion among low-income veterans in our study is consistent with broader evidence that poverty amplifies digital access and usage gaps [37]. Intersectional approaches are therefore needed when designing digital inclusion programmes.

Our findings match previous research on digital exclusion and mental health. A survey follow-up in mental health services found that online surveys overestimated digital inclusion because individuals without internet access could not participate; digital exclusion persisted despite modest improvements and was highest among people with chronic psychosis and older adults [33]. These results underscore studies’ reliance on online recruitment, which may underestimate the scale of exclusion, a key limitation of the current study. Qualitative studies have described digital exclusion among mental health service users, highlighting barriers such as lack of knowledge, material deprivation, rural isolation and mental health symptoms [35].

Our quantitative data reinforce these themes, showing that self-reported digital exclusion is linked to cost barriers, privacy concerns and difficulties using technology. Research on adolescents during the Covid-19 pandemic found that lacking a computer predicted poorer mental health trajectories [39], suggesting that digital access is increasingly important across the life course.

Key strengths of the study include the large sample of UK veterans surveyed and the use of validated screeners for depression, anxiety and loneliness. We adapted measures from a US veterans’ digital needs survey, enabling international comparison and use of a consistent digital exclusion question. We also captured multiple facets of digital inclusion (access, confidence, cost barriers, privacy concerns), allowing analysis of the digital divide.

This study has several limitations. First, the cross-sectional design precludes causal inference. Depressive symptoms, anxiety or loneliness may contribute to digital disengagement, just as digital exclusion may exacerbate difficulties accessing services, information and social support. Longitudinal studies are needed to disentangle these pathways. Second, all data were self-reported and may be affected by recall bias, social desirability bias and misclassification. Third, the survey was conducted online and therefore required participants to have some degree of digital access, confidence or support. Veterans experiencing the most severe forms of digital exclusion, including those without internet access or a suitable device, are therefore likely to be underrepresented. As a result, the prevalence of digital exclusion reported here should not be interpreted as a definitive population estimate for all UK veterans. Fourth, the primary exposure was based on a single self-report item capturing perceived exclusion or disadvantage due to lack of digital access or skills. This approach is useful for identifying subjective disadvantage, but it does not capture the full complexity of digital exclusion, including objective access, affordability, skills, motivation, trust and service design. Future studies should combine subjective and objective measures of digital inclusion and consider mixed methods approaches.

Our findings have implications for efforts to adopt a ‘digital first’ approach in health and public services. The association between digital exclusion and poorer mental health suggests that digital inequalities could widen existing health disparities. Clinicians and service providers should routinely assess patients’ digital access and skills and offer tailored support. This could include the loaning of devices with training and support. Evidence from mental health services indicates that training and social support can facilitate engagement with digital interventions [34], while anonymity and privacy can be both facilitators and barriers. Policy makers should invest in targeted programmes that address affordability, skills (hands-on training, helplines) and accessibility (simple user interfaces, language options). Programmes should be inclusive of veterans and consider intersections with age, gender, income and health status. This also could include occupational health programmes where use of digital technology and generative AI is increasing [2]. Importantly, alternative non-digital routes to care must remain available; universal digital requirements could inadvertently exclude those most in need [40].

## Conclusion

More than two-fifths of UK veterans with valid exposure data in this online survey reported feeling digitally excluded or disadvantaged, despite high levels of reported device access and confidence. Self-reported digital exclusion was associated with higher odds of probable depression, probable anxiety and probable loneliness after adjustment for sociodemographic and service-related factors. These findings suggest that the key digital divide for many veterans may not be access alone, but whether digital services are affordable, trusted, usable and capable of meeting their needs. Because the study was conducted online, veterans with the most severe digital exclusion are likely to be underrepresented. Digital-first services for veterans should therefore include routine digital needs screening, targeted support, clear privacy reassurance and accessible non-digital routes to care.

## Data Availability

All data produced in the present study are available upon reasonable request to the authors

## Data availability

The dataset supporting the findings of this study will be made available via the Open Science Framework (<DOI upon acceptance>).

